# FACTORS INFLUENCING CONTRACEPTIVE USE AMONG ADOLESCENTS IN TECHIMAN MUNICIPALITY, GHANA

**DOI:** 10.1101/2022.07.29.22278209

**Authors:** Titus N. Kpiinfaar, Gerald Owusu-Asubonteng, Edward T. Dassah

## Abstract

Contraceptive use among adolescents in Ghana is low. Factors influencing adolescent contraceptive use in Techiman Municipality have not been investigated. This study aimed to determine factors influencing contraceptive use among adolescents in Techiman Municipality.

An analytical cross-sectional community-based study was conducted from January-March, 2018 among adolescents between the ages of 16 and 19 years whoever had sex. Respondents were selected through multi-stage sampling. Data was collected using a pre-tested structured questionnaire. Factors associated with contraceptive use were assessed using logistic regression to generate crude and adjusted odds ratios (AORs) with 95% confidence intervals (CIs). P<0.05 was considered statistically significant. Altogether, 298 adolescents who have ever had sex were studied. The mean age was 17.5 years (standard deviation 1.1 years). About 53% were out-of-school adolescents and 43.6% had sex within the four weeks preceding the interview, with out-of-school adolescents being more likely to have had sex (53.8% vs. 32.1%; p<0.001). Contraceptive knowledge and ever use were 75.0% and 65% respectively. The most commonly used contraceptives were condoms (54%) and emergency contraception (31%). Significant factors influencing contraceptive use were; father’s educational level (AOR4.86; 95% CI 1.70 – 13.91; p<0.001) and prior discussion of contraceptive use with partner (AOR 3.96; 95% CI 1.32 – 11.89; p=0.01). Contraceptive use was high, but few (10%) relied on LARCs. Adolescents should be encouraged to discuss contraceptive use with their sexual partners during sexual health and contraceptive counselling. Strategies for improving LARC uptake among adolescents should be investigated.

## INTRODUCTION

Globally, prevention of adolescent pregnancy and early childbirth are critical healthcare issues [1]. About 21million adolescent pregnancies are recorded worldwide each year, and almost half (49%) of pregnancies are unintended, and a little over half result in induced abortions (51%) [1]. Nearly a quarter (23%) of all adolescent births occurs in Africa [3]. Adolescent pregnancies occur in areas with low contraceptive prevalence [4]. Sub-Saharan Africa has the lowest contraceptive demand (30%) and use (20%) among adolescents [5]. The commonly used modern contraceptives methods by adolescents in Africa include, male condoms (38%) [2], emergency contraceptive pill (27%), injectables (19%), implants (8%) and intra-uterine devices (IUDs 5%) [6]. The low contraceptive uptake results in teenage pregnancies, high childbirth rates and incidence of sexually transmitted infections (STIs) including HIV [7].

Only about 8% of married adolescents in Ghana use modern contraceptives but the rates vary across the country [8]. Contraceptive use among the general adolescent population may even be lower. For example, contraceptive use among adolescents in Techiman Municipality in 2016 was 1.6% with a correspondingly high teenage pregnancy rate of 17.1%. A number of studies have investigated contraceptive use in the general population [9]. However, there is limited data on factors associated with contraceptive use among adolescents in Ghana especially in Techiman Municipality. This study determined factors influencing contraceptive use among adolescents in Techiman Municipality.

## MATERIALS AND METHODS

### Study design

This was an analytical cross-sectional community-based survey conducted among adolescents in Techiman Municipality from January to March, 2018. Aadolescents between the ages of 16 and 19 years who ever had sex and had been residing in the municipality for at least 6 months prior to the study and gave consent to participate in the study were eligible for inclusion.

### Study setting

Techiman Municipality is one of the 27 administrative districts in the then Brong Ahafo Region. The municipality is currently regional capital of the newly created Bono East Region. The municipality has seven sub-municipals, with its capital at Techiman, a commercial town with about 64.5% of its citizens living in urban areas. The municipality had a projected population of 147,788 people. About half (51.5%) of the population were females with 39,718 (26.9%) being in their reproductive ages (15-49 years). Average household size was 4 persons per household [10].

### Study procedures

Participants for the study were selected through multistage sampling. The list of communities and number of adolescents within each sub-municipal was obtained from the Municipal Assembly. Four of the seven sub-municipals in the Techiman municipal were selected by simple random sampling through balloting; Tanoso, Techiman-West, Techiman-North and Nkwaeso. The number of communities within the selected sub-municipals varied from 14-22. The number of adolescents recruited from each sub-municipal was obtained in proportion to the total number of adolescents in the chosen sub-municipals. Within each sub-municipal, one community was selected by simple random sampling [11].

In each selected community a central reference point such as a borehole, church or mosque was identified. Here, a pen was span and every other household in the direction of the pen was chosen [11]. Where the selected household did not have an eligible adolescent; the next household in the direction visited was selected. After reaching the last household in a particular direction, we returned to the central point and the procedure was repeated in a different direction until the desired sample size for the community was attained. At the household level the study was explained to the household, and permission obtained from the head. Within each household, resident adolescents between the ages of 16-19 years were invited individually by a member of the research team to a private place to enquire about sexual activity and those who met the criteria were invited to participate. Where there was more than one eligible adolescent, only one was randomly selected through balloting.

Selected adolescents were invited for interview at a suitable place within or outside the house. After explaining the purposes of the study, informed consent was obtained and a confidential interview conducted in English or a local language understood by both the research team and the participant. In the case of adolescents under 18 years, consent was obtained from a parent/guardian with assent from the adolescent. Using a structured questionnaire, data were collected on their socio-demographic characteristics, knowledge of contraceptive methods and factors influencing contraceptive use. Pretesting of the questionnaire was done in a sub-municipal (Forikrom) which was not part of the selected sub-municipals.

The main outcome in this study was contraceptive use. Contraceptive use was defined as the intentional prevention of pregnancy (conception) through the use of various contraceptive devices like male and female condoms, injectables, Intra-Uterine Device (IUD), implants. Sexual experience and contraceptive use were self-reported by respondents. Adolescent’s knowledge of contraceptive use was assessed by evaluating their responses to ten questions on contraceptive use including; awareness, sources of awareness, method use, reason and contraceptive side effects. Each correct response attracted a score of “+1” while each “incorrect” or “undecided” (“don’t know”) response was assigned a score of “0”. The scores for each adolescent were summed and graded as follows; scores 0-3 methods = poor knowledge, 4-6 methods = average knowledge and 7-10 methods = good knowledge.

### Sample size calculation and statistical analyses

Sample size calculations were done in Epi Info STATCALC version 7 with 95% confidence level and a power of 80% and a margin of error of 5% [11]. Assuming the factors influencing contraceptive use were similar to those observed by Boamah et al in Kintampo [3], and allowing for 10% contingency, an estimated sample size of 300 was adequate to determine these factors. Data was cross-checked for completeness before double-entry into an excel database.

Data was cleaned and exported into STATA version 12.1 (Stata Corp, College Station, Texas, USA) for analysis. Categorical and continuous variables were compared using the chi-square test and student t-test respectively. Factors associated with contraceptive use were assessed using logistic regression to generate odds ratios (ORs) with 95% confidence intervals (CIs). Statistical significance was set at p ≤ 0.05. All missing values were excluded from the analyses.

### Ethics approval

The study was conducted in accordance with the Declaration of Helsinki and was approved by the Committee on Human Research, Publications and Ethics of the School of Medical Science, Kwame Nkrumah University of Science and Technology, Kumasi (CHRPE/AP/434/17).

Participants gave informed consent to participate in the study before they were recruited into the study. For adolescents under 18 years, consent was obtained from a parent/guardian with assent from the adolescent.

## RESULTS

A total of 300 adolescents were invited to participate, of whom two declined consent and the remaining 298 were recruited into the study. Table 1 summarizes the socio-demographic characteristics of respondents. The mean age was 17.5 years (standard deviation 1.1 years), and range 16-18 years. They were virtually distributed equally between the two age groups, half (50%) were females and same were males. More than half (53.1%) were out-of-school adolescents and 65% had completed basic education. About 5.7% were Akans and 74.5% were Christians. Over 90% were single and about a third each lived with both parents, either parent, a relative/guardian or in a rural area. About three in 10 were engaged in some form of employment. Over 70% of their fathers had attained some level of formal education.

**Table 1:** Socio-demographic characteristics of the adolescents.

| **Table1: Socio-demographic characteristics of the adolescents Variable** | **Frequency (N=298)** | **Percentage (%)** |
| --- | --- | --- |
| Age group (years) |  |  |
| 16-17 | 148 | 49.7 |
| 18-19 | 150 | 50.3 |
| Sex |  |  |
| Male | 149 | 50.0 |
| Female | 149 | 50.0 |
| Current educational status |  |  |
| In-school | 140 | 46.9 |
| Out-of-school | 158 | 53.1 |
| Respondents' educational level |  |  |
| No formal education | 14 | 4.7 |
| Basic education | 193 | 64.8 |
| SHS and above | 91 | 30.5 |
| Ethnic group |  |  |
| Akan | 169 | 56.7 |
| Mole-Dagbani | 114 | 38.3 |
| Other tribes | 15 | 5.1 |
| Religion |  |  |
| Christianity | 222 | 74.5 |
| Islam/Traditional | 76 | 25.5 |
| Marital status |  |  |
| Single | 278 | 93.3 |
| Married | 20 | 6.7 |
| Living arrangement |  |  |
| Both parents | 105 | 35.2 |
| Either parent | 92 | 30.9 |
| Husband/Other relatives | 101 | 33.9 |
| Residential status |  |  |
| Rural | 95 | 31.9 |
| Urban | 203 | 68.1 |
| Occupational status |  |  |
| Unemployed | 207 | 69.5 |
| Employed | 91 | 30.5 |
| Mother's educational level |  |  |
| No formal education | 79 | 26.5 |
| Basic education | 110 | 36.9 |
| SHS and above | 109 | 36.6 |
| Father's educational level |  |  |
| No formal education | 85 | 28.5 |
| Basic education | 105 | 35.3 |
| SHS and above | 108 | 36.2 |

Adolescents’ knowledge of contraceptive methods is summarized in Table 2 below. Nearly two-thirds (66.1%) had knowledge of at least three contraceptive methods, 29.2% had knowledge of 4-6 contraceptive methods and only 5% had knowledge of up to ten contraceptive methods. Three-quarters (75%) of sexually experienced adolescents had knowledge of method before use, and 64.6% of their sexual partners were reported to have had knowledge of method prior to its use. Sources of information on contraceptives included: school/teacher (37.8%), friends/partners (28.8%), electronic/print media (13.5%), health workers (11.2%) and parents/relatives (8.6%). Forty-four percent had had sex within the four weeks preceding the interview, out-of-school adolescents were more likely to be sexually experienced (53.8% vs. 32.1%; p<0.001). More than half (56.4%) had had no sexual activity within the three months preceding the survey, and 28.5% were coerced/forced into having sex.

**Table 2:**
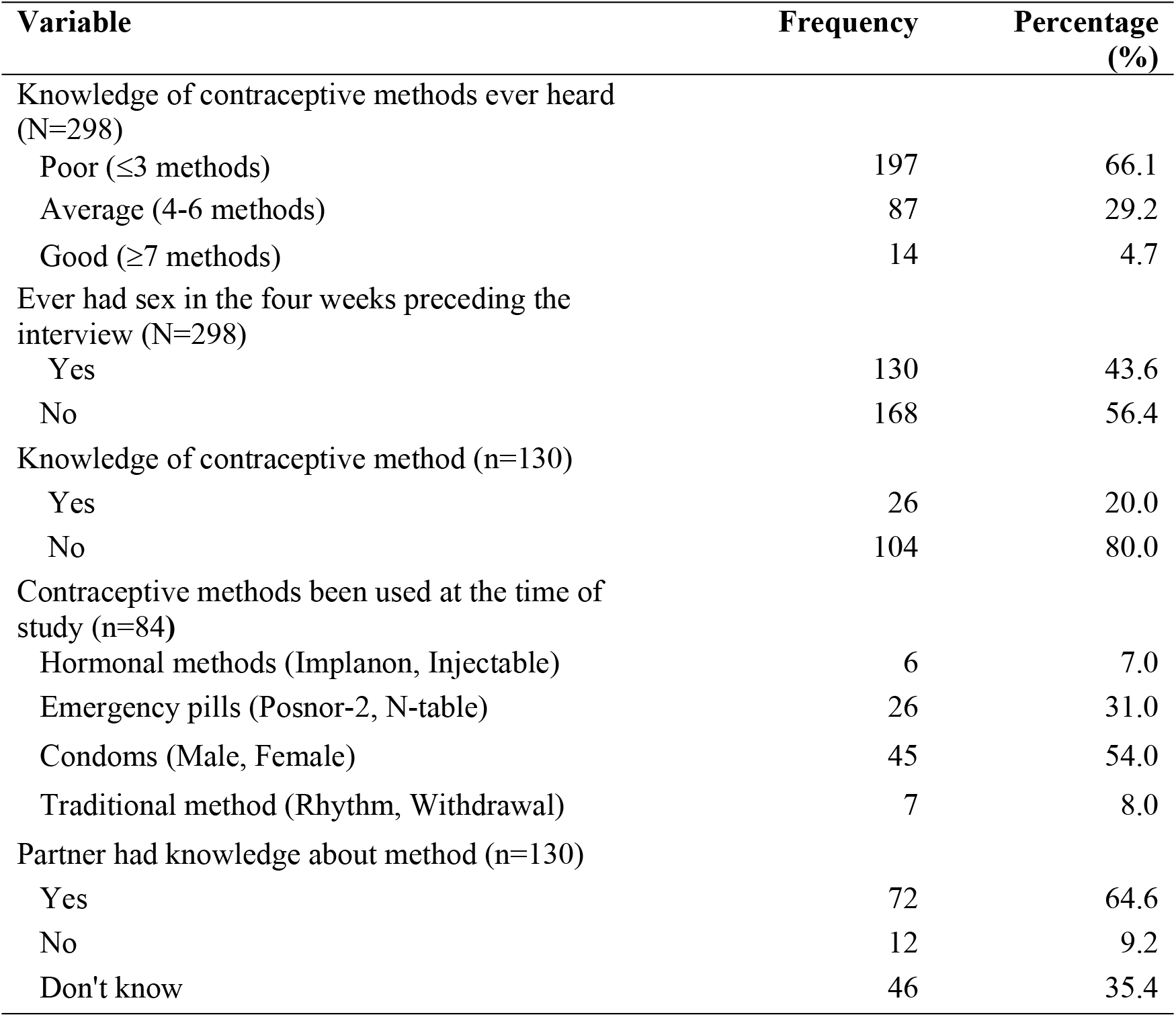
Knowledge of contraceptives among adolescents.

| Variable | Frequency | Percentage (%) |
| --- | --- | --- |
| Knowledge of contraceptive methods ever heard (N=298) |  |  |
| Poor ($\leq 3$ methods) | 197 | 66.1 |
| Average (4-6 methods) | 87 | 29.2 |
| Good ($\geq 7$ methods) | 14 | 4.7 |
| Ever had sex in the four weeks preceding the interview (N=298) |  |  |
| Yes | 130 | 43.6 |
| No | 168 | 56.4 |
| Knowledge of contraceptive method (n=130) |  |  |
| Yes | 26 | 20.0 |
| No | 104 | 80.0 |
| Contraceptive methods been used at the time of study (n=84) |  |  |
| Hormonal methods (Implanon, Injectable) | 6 | 7.0 |
| Emergency pills (Posnor-2, N-table) | 26 | 31.0 |
| Condoms (Male, Female) | 45 | 54.0 |
| Traditional method (Rhythm, Withdrawal) | 7 | 8.0 |
| Partner had knowledge about method (n=130) |  |  |
| Yes | 72 | 64.6 |
| No | 12 | 9.2 |
| Don't know | 46 | 35.4 |

Overall, 64.6% of adolescents whoever had sex had ever used contraceptives. The commonly used contraceptive methods were: condoms (54%), emergency pills (31.0%), injectables and implants (7%) and traditional methods such as rhythm and withdrawal methods (8%). Less than 10% were using long-acting reversible contraceptive (LARC) methods. The decision to use contraceptives was taken together with their partners (49.5%), solely by their partners (41%) or with their parents/friends (9.5%). About 72% found it difficult to discuss contraceptive use with their partners during their first sex because they felt ashamed to talk of contraceptives, while 28% had discussed contraceptive use with their partner before the first sexual encounter. About 30% of parents discussed sexual issues with their adolescent girls while 70% never did because they felt their adolescent girls were still young. On future contraceptive intentions, 65% had intentions of using contraceptives in the future.

Factors influencing ever use of contraceptives are shown in Table 3 below. On univariable analysis, significant factors associated with ever use of contraceptives were; the adolescent’s or father’s educational level and prior discussion of contraceptive use with partner. On multivariable analysis, the educational level of the father and prior discussion of contraceptive use with partner remained as independent predictors of contraceptive use. Adolescents whose fathers had had no formal education (adjusted OR, 3.85; 95% CI 1.46-10.17) or at least secondary education (adjusted OR 4.86; 95% CI 1.70-13.91) were more likely to have used contraceptives compared to those whose fathers had attained basic education. Discussion of contraceptive use with partner prior to sexual intercourse increased the odds of contraceptive use by nearly four-fold (adjusted OR 3.96; 95% CI1.32-11.89).

**Table 3:** Univariable and multivariable analysis of factors influencing contraceptive use among adolescents in Techiman Municipality, Ghana.

| Variable | Ever use of<br>contraceptives<br>n (%) | Unadjusted OR<br>(95% CI) | Adjusted OR<br>(95% CI) |
| --- | --- | --- | --- |
| Age group (years) |  | p=0.60 | - |
| 16-17 | 29 (61.7) | 1 | - |
| 18-19 | 55 (66.3) | 1.22 (0.58-2.56) | - |
| Current educational status |  | p=0.06 | - |
| Out-of-school | 50 (58.8) | 1 | - |
| In-school | 34 (75.6) | 0.46 (0.20-1.04) | - |
| Educational level of adolescent |  | p=0.01 | p=0.08 |
| No formal education | 3 (42.9) | 1 | 1 |
| Basic education | 49 (57.7) | 7.11 (0.08-46.52) | 4.38 (0.64-29.84) |
| SHS and above | 32 (84.2) | 1.81 (0.38-8.73) | 1.35 (0.25-7.25) |
| Mother's educational level |  | p = 0.25 |  |
| No formal education | 34 (61.8) | 1 | - |
| Basic education | 18 (75.0) | 1.85(0.62-5.49) | - |
| SHS and above | 32 (62.8) | 1.04 (0.47-2.29) | - |
| Father's educational level |  | p=0.003 | p=0.02 |
| No formal education | 27 (71.1) | 1 | 1 |
| Basic education | 22 (44.0) | 0.32 (0.13-0.82) | 3.85 (1.45-10.17) |
| SHS and above | 35 (83.3) | 2.04 (0.68-6.06) | 4.86 (1.70-13.91) |
| Able to discuss sex issues with<br>parents |  | p=0.06 | - |
| No | 52 (59.8) | 1 | - |
| Yes | 32 (76.2) | 2.15 (0.94-4.93) | - |
| Ever discussed contraceptive use<br>with partner before sex |  | p=0.01 | p=0.01 |
| No | 53 (56.4) | 1 | 1 |
| Yes | 31 (86.1) | 4.79 (1.71- 13.42) | 3.96 (1.32- 11.89) |

## DISCUSSION

This study sought to determine factors influencing contraceptive use among adolescents in Techiman municipality. Only about a third of these adolescents had average to good knowledge of contraceptive methods. Majority had ever used contraceptives but less than a tenth relied on LARCs. Significant factors associated with contraceptive use were father’s educational background and prior discussion of contraceptive use with partner.

About one-third of the adolescents had good knowledge of contraceptive methods. Similarly, in Nigeria 75% of adolescents were found to have good knowledge of contraceptive methods [12]. However, the proportion of adolescents with contraceptive knowledge in our study was much higher than the 17% reported in nationwide survey [8 - 16]. The higher proportion in our study could be due to improved health education on contraceptive use in schools and among the general public.

Contraceptive use among adolescents in the municipality was quite high as compared to a previous national survey rate of 19% [8]. The higher contraceptive uptake in our study could be attributed to the provision of free contraceptive services to adolescents in the municipality by non-governmental organizations such as the West Africa Program to Combat AIDS and STI (WAPCAS) and Planned Parenthood Association of Ghana (PPAG) [13]. These findings may suggest that in-school adolescents in the municipality could be at a lower risk of teenage pregnancy provided the contraceptives are being used correctly and consistently. The commonly used contraceptives among adolescents in the current study have been documented in previous studies [14]. However, it is quite worrying that a vast majority (>80%) relied non-LARC methods (mainly condoms and emergency contraceptives), which are less effective contraceptives as compared to LARCs [18]. While condoms have the advantage of offering dual protection (against pregnancy and STIs) they are less efficacious contraceptives than LARCs [15]. This possibly explains the relatively high teenage pregnancy rate despite the contraceptive uptake among adolescents in the municipality.

Adolescents with formal educational background were more likely to use contraceptives as compared to those without any form of formal education. This could be due to exposure tocontraceptive information in school. These findings are consistent with those of previous studies in Ghana [19] and Nigeria [20]. As observed in Nigeria, contraceptive use increased with increased educational status of the girls’ fathers. Increasing educational status enhances access to health services and access to information on various Reproductive Health issues [12 - 23]. It also increases access to employment and disposable income, which has been shown to be a predictor of contraceptive use in adolescents [17]. It is also possible the adolescents were adequately exposed to Sexual Reproductive Health (SRH) information in their school as SRH is taught in Ghanaian schools [19 - 22]. Prior discussion of contraceptives use among adolescents who ever had sex before sexual activity promote contraceptive use among adolescent as it was found to have had four-fold increase in the likelihood of using contraceptives when compared with those who had never discussed about contraceptives before sex [24 - 25]. Promoting such discussion before sex ensures mutual engagement and understanding among adolescents on the choice of contraceptive method to use [21 - 26].

### Strength and Limitations

A major strength of this study was that being a population-based study that employed multistage sampling, the study population was representative of the adolescent population in the study area. However, the study had some limitations. Adolescents who were not at home during the study were excluded. Their sexual and contraceptive experiences could well be different from those that were at home. Sexual activity and contraceptive use which key outcomes in the study are sensitive information. These were self-reported and could be subject to reporting bias. Participants were encouraged to be as candid as possible to minimize bias.

### Conclusion

Contraceptive use among adolescents was high,but a vast majority relied on non-LARC methods. Discussing contraceptive use with sexual partners should be encouraged during sexual health and contraceptive counselling, while strategies for improving LARC uptake among adolescents in the municipality are investigated.

## Data Availability

Data relating to the study findings has been attached as "Supporting Information" for your attention. Database on the factors influencing contraceptive use among adolescents in Techiman Municipality, 2018. Variables: id number, age, sex, level of education, religion, marital status, living arrangement, residential status, ever use contraceptive (yes = 1, no = 2), current use of contraceptives (yes = 1, no = 2), ever had sex (yes = 1, no = 2), current sex (yes = 1, no = 2), aware of contraceptive method before use (yes = 1, no = 2).

## Supporting information

**S1 Data. Database on the factors influencing contraceptive use among adolescents in Techiman Municipality, 2018**. Variables: id number, age, sex, level of education, religion, marital status, living arrangement, residential status, ever use contraceptive (yes = 1, no = 2), current use of contraceptives (yes = 1, no = 2), ever had sex (yes = 1, no = 2), current sex (yes = 1, no = 2), aware of contraceptive method before use (yes = 1, no = 2).

## Acknowledgement

We are very grateful to the entire staff of Techiman Municipal Health Directorate, the Administrator of Holy Family Hospital (Techiman), and the School of Public Health (KNUST).

## Author Contributions

**Conceptualization:** Titus N. Kpiinfar, Gerald Owusu-Asubonteng, Edward T. Dassah

**Data curation:** Titus N. Kpiinfar

**Formal analysis:** Titus N. Kpiinfar, Edward T. Dassah

**Funding acquisition:** Titus N Kpiinfaar

**Methodology:** Titus N. Kpiinfar, Gerald Owusu-Asubonteng, Edward T. Dassah

**Resources:** Titus N. Kpiinfar

**Supervision:** Edward T. Dassah

**Validation:** Titus N. Kpiinfar, Edward T. Dassah

**Writing – original draft:** Titus N. Kpiinfar, Edward T. Dassah

**Writing – review & editing:** Titus N. Kpiinfar, Gerald Owusu-Asubonteng, Edward T. Dassah

**Funding:** The authors received no specific funding for this work.

**Competing interests**: The authors have declared that no competing interests exist.

